# Automated Pediatric Brain Tumor Imaging Assessment Tool from CBTN: Enhancing Suprasellar Region Inclusion and Managing Limited Data with Deep Learning

**DOI:** 10.1101/2024.07.29.24311006

**Authors:** Deep B. Gandhi, Nastaran Khalili, Ariana M. Familiar, Anurag Gottipati, Neda Khalili, Wenxin Tu, Shuvanjan Haldar, Hannah Anderson, Karthik Viswanathan, Phillip B. Storm, Jeffrey B. Ware, Adam Resnick, Arastoo Vossough, Ali Nabavizadeh, Anahita Fathi Kazerooni

## Abstract

**Background:** Fully-automatic skull-stripping and tumor segmentation are crucial for monitoring pediatric brain tumors (PBT). Current methods, however, often lack generalizability, particularly for rare tumors in the sellar/suprasellar regions and when applied to real-world clinical data in limited data scenarios. To address these challenges, we propose AI-driven techniques for skull-stripping and tumor segmentation.

**Methods:** Multi-institutional, multi-parametric MRI scans from 527 pediatric patients (n=336 for skull-stripping, n=489 for tumor segmentation) with various PBT histologies were processed to train separate nnU-Net-based deep learning models for skull-stripping, whole tumor (WT), and enhancing tumor (ET) segmentation. These models utilized single (T2/FLAIR) or multiple (T1-Gd and T2/FLAIR) input imaging sequences. Performance was evaluated using Dice scores, sensitivity, and 95% Hausdorff distances. Statistical comparisons included paired or unpaired two-sample t-tests and Pearson’s correlation coefficient based on Dice scores from different models and PBT histologies.

**Results:** Dice scores for the skull-stripping models for whole brain and sellar/suprasellar region segmentation were 0.98±0.01 (median 0.98) for both multi- and single-parametric models, with significant Pearson’s correlation coefficient between single- and multi-parametric Dice scores (r > 0.80; p<0.05 for all). WT Dice scores for single-input tumor segmentation models were 0.84±0.17 (median=0.90) for T2 and 0.82±0.19 (median=0.89) for FLAIR inputs. ET Dice scores were 0.65±0.35 (median=0.79) for T1-Gd+FLAIR and 0.64±0.36 (median=0.79) for T1-Gd+T2 inputs.

**Conclusion:** Our skull-stripping models demonstrate excellent performance and include sellar/suprasellar regions, using single- or multi-parametric inputs. Additionally, our automated tumor segmentation models can reliably delineate whole lesions and enhancing tumor regions, adapting to MRI sessions with missing sequences in limited data context.

**Brief key points:**

1. Deep learning models for skull-stripping, including the sellar/suprasellar regions, demonstrate robustness across various pediatric brain tumor histologies.
2. The automated brain tumor segmentation models perform reliably even in limited data scenarios.

**Importance of the Study:** We present robust skull-stripping models that work with single- and multi-parametric MR images and include the sellar-suprasellar regions in the extracted brain tissue. Since ∼10% of the pediatric brain tumors originate in the sellar/suprasellar region, including the deep-seated regions within the extracted brain tissue makes these models generalizable for a wider range of tumor histologies. We also present two tumor segmentation models, one for segmenting whole tumor using T2/FLAIR images, and another for segmenting enhancing tumor region using T1-Gd and T2/FLAIR images. These models demonstrate excellent performance with limited input. Both the skull-stripping and tumor segmentation models work with one- or two-input MRI sequences, making them useful in cases where multi-parametric images are not available – especially in real-world clinical scenarios. These models help to address the issue of missing data, making it possible to include subjects for longitudinal assessment and monitoring treatment response, which would have otherwise been excluded.

## INTRODUCTION

Pediatric brain tumors (PBTs) are the most prevalent childhood cancers of the central nervous system (CNS), encompassing a wide range of histologies and survival rates ^1–4^, and are one of the leading causes of cancer-related deaths in children, only secondary to leaukemia ^5–7^. The World Health Organization (WHO), in the 5^th^ edition of its Classification of Tumors of the Central Nervous System (WHO CNS5), recognizes that PBTs possess distinct histological and molecular features ^8^. Consequently, there are notable differences in neuroimaging characteristics between adult and pediatric brain tumors, including variations in brain structures, image signal intensity, skull formation, and tumor subregions ^9^. These differences underscore the need for image processing and assessment tools tailored specifically to pediatric neuroimaging data.

Quantitative analysis of PBTs for response assessment requires accurately locating and delineating the tumorous region, a challenging and tedious task prone to inter-reader variability and lack of consensus ^10,11^. While established automated preprocessing and tumor size measurement approaches exist for adult brain tumors ^12^, and despite recent advances in developing pediatric-specific automated methods for tumor assessment ^13–18^, there still remains a lack of comprehensive methods addressing the unique challenges of tumor assessment in pediatric patients.

Automated pediatric-specific approaches employing deep learning models, such as Convolutional Neural Networks (CNNs), have been utilized for skull-stripping and segmentation of whole lesion or tumor subregions ^9,14–16,19^. Skull-stripping, also referred to as brain extraction, is a crucial image pre-processing technique for isolating brain tissue from non-brain tissue in MRI. This step is vital for downstream neuroimaging analyses and plays an essential role in ensuring patient anonymization during data sharing. Various imaging analysis methodologies, including image intensity standardization for radiomic feature extraction, image registration, tumor segmentation, and the mapping of MRI to other imaging modalities, achieve higher accuracy when the images are skull-stripped ^20,21^.

Previous skull-stripping methods were either developed using MRI data from patients without brain tumors ^22^, based on adult brain tumors ^20,23^, or, although trained for PBTs – as in our previous study ^16^ – did not adequately cover deep-seated brain regions such as the sellar/suprasellar areas, leading to undersegmentation of tumors in these regions. In pediatrics, sellar and suprasellar tumors account for approximately 10% of all CNS tumors and encompass a diverse array of entities, each with unique histologic origins and radiological features ^24^. These tumors often present with specific clinical and neuroimaging characteristics, necessitating tailored surgical interventions and therapeutic approaches ^24^. Therefore, it is essential to accurately include these regions within the brain tissue for image processing tools to be generalizable across various PBT histologies. Furthermore, given that sellar/suprasellar tumors can distort the anatomy of the optic pathway, it is crucial to develop a tool that improves the extraction of brain tissue while preserving the sellar/suprasellar region ^25^.

In our earlier work ^15,16^, we developed multi-parametric tumor subregion segmentation models capable of efficiently predicting the whole tumor (WT) and different tumor subregions, including enhancing tumor (ET) core, non-enhancing tumor (NET) core, cystic component (CC), and peritumoral edema (ED), for a variety of pediatric brain tumors using four standard MRI sequences: T1-weighted (T1), T1-weighted post-contrast enhanced (T1-Gd), T2-weighted (T2), and T2-weighted fluid attenuated inversion recovery (FLAIR) images. However, in some instances, depending on the purpose of the imaging (e.g., initial assessment versus follow-up imaging), not all four sequences are acquired at a given timepoint, or the images may be unusable due to artifacts or specific protocol settings. This lack of availability of multi-parametric scans, particularly in retrospective studies and longitudinal tumor response assessments, is especially pronounced in pediatric cases. The relatively low incidence of brain tumors in the pediatric population necessitates data collection from multiple sites, each with differing clinical protocols, leading to decreased harmonization of input data ^26^.

This inconsistency may result in the exclusion of subjects who otherwise meet eligibility criteria and could be included for model training and further analysis. Nevertheless, based on tumor histology, single-parametric scans can still provide valuable clinical information. For instance, in the context of diffuse midline glioma (DMG), delineating the whole tumor based on T2 and/or FLAIR scans may be sufficient for longitudinal response assessment ^27^. In the case of tumors with enhancing components, such as pediatric low-grade and high-grade gliomas (LGG and HGG; respectively), segmentation of the enhancing tumor based on T1-Gd images could be beneficial for evaluating tumor behavior and likely progression ^28,29^. Although several studies have explored the efficacy of single versus multi-parametric MRI for developing segmentation pipelines in CNS lesions such as meningioma or vestibular schwannoma ^30,31^, no established pre-processing pipelines or tools currently exist to address this problem in PBTs.

To address these unmet needs, we propose a generalizable pediatric pre-processing pipeline for enhanced automated brain tissue extraction (skull-stripping) and tumor segmentation. This pipeline complements our previously developed multi-parametric auto skull-stripping and tumor segmentation models. In this study, we trained 3D convolutional neural networks (CNNs) using a U-Net-based architecture (nnU-Net) ^32^ with multi-parametric and single-parametric MRI sequences as inputs for the auto-segmentation tasks. We selected nnU-Net because it has been proven to outperform most available CNNs, especially in applications involving PBTs ^15,33,34^.

We hypothesize that this pipeline can establish a standardized method for pre-processing pediatric brain MRI acquisitions. Furthermore, we expect that the proposed auto-segmentation models will demonstrate acceptable performance in segmenting either the WT or ET components, including sellar and suprasellar regions, even in subjects lacking multi-parametric MRI scans.

## METHODS

### Data description and Patient cohort

Retrospective data from pediatric subjects was collected from the multi-institutional Children’s Brain Tumor Network (CBTN) repository and BraTS-PEDs 2023 ^35,36^. Subjects were included if they had the following four MR images obtained routinely for clinical evaluation of brain tumors: T1, T1-Gd, T2 and FLAIR. Additionally, only subjects who underwent minor surgical procedures that did not result in major neuroanatomical changes and had all four brain MR images mentioned above, were included. Subjects were excluded if they they underwent surgical procedures that resulted in major changes to neuroanatomy.

Based on the availability of ground truth brain masks and tumor segmentations, two different subject cohorts were created as a part of this study, one for training the skull-stripping models and another for training the tumor segmentation models. Detailed description of the patient demographics and distribution of tumor histology for each cohort is provided (Figure 1).

**Figure 1:**
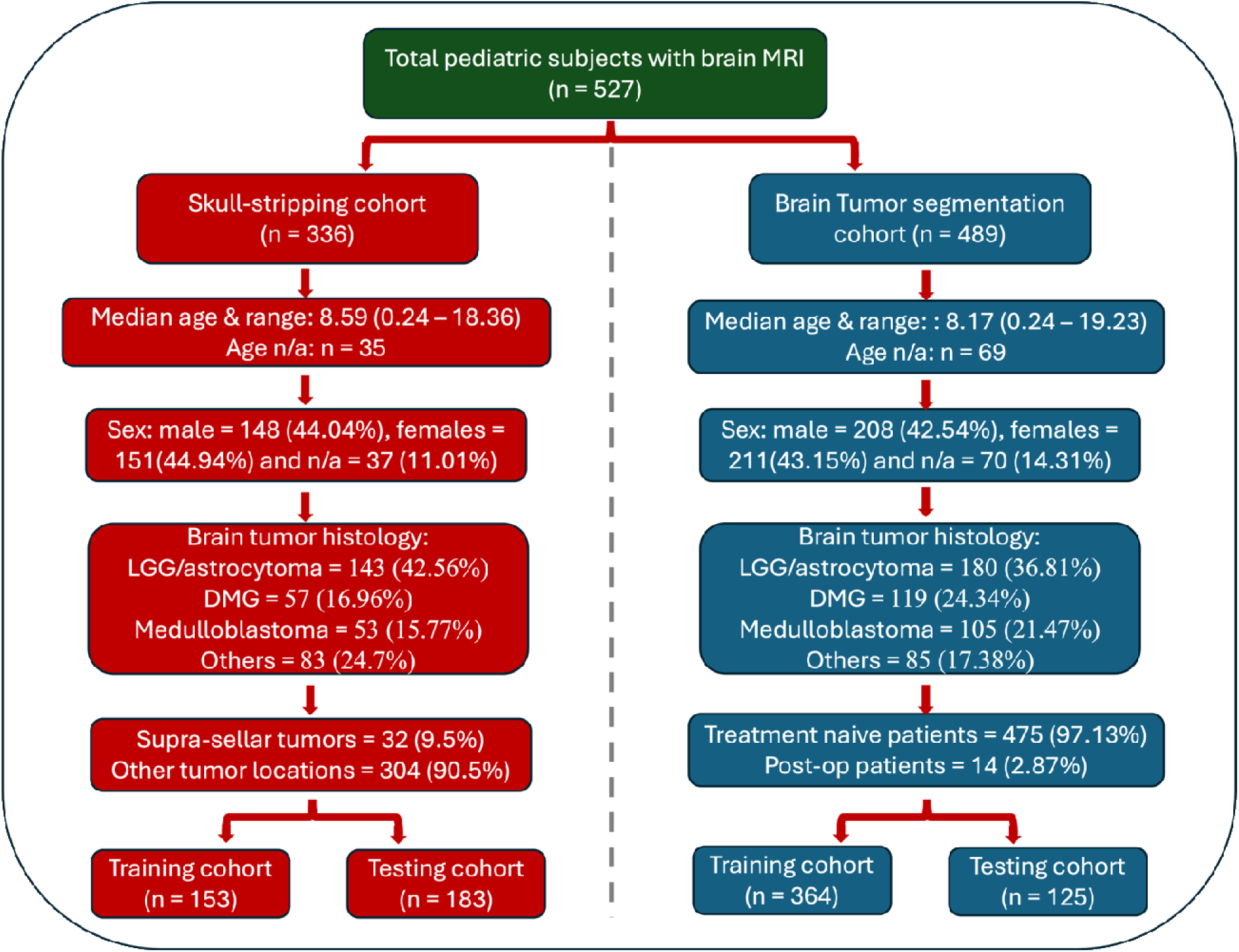
Patient demographics for subjects used in model training and testing for the skull-stripping and brain tumor segmentation cohorts.

### Image pre-processing and data preparation

Data preparation for single and multi-parametric model training included a series of pre-processing steps (Supplemental Figure 1). First, all images were reoriented to left-posterior-superior (LPS) co-ordinate system. Next, the T1-Gd image was co-registered to the SRI24-atlas space and subsequently the T1, T2 and FLAIR images were co-registered to the T1-Gd image. Images were resampled to 1 mm^3^ isotropic resolution and the image dimensions were changed to 240×240×155, based on the anatomical SR124-atlas space ^37^. Co-registration was performed using a greedy algorithm in the Cancer Imaging Phenomics Toolkit open-source software v.1.8.1 (CaPTk, https://www.cbica.upenn.edu/captk) ^38,39^.

A semi-automated process was used to create ground truth segmentation masks for model training. For the skull-stripping model, images were passed through an existing automated skull-stripping tool based on DeepMedic from CaPTk ^40^. The resulting brain masks underwent manual modification to make any corrections and, importantly, to include the sellar/suprasellar regions within the brain masks. Similarly, to generate initial tumor segmentations, images were passed through a baseline automated tumor segmentation tool ^15^, which segmented four various tumor subregions – ET, NET, CC, and ED - where present. For the baseline segmentations generated above, manual revisions were made by trained researchers using ITK-SNAP ^41^. These revisions were then reviewed by one of the three practicing neuroradiologists (J.W. with 6, A.N. with 10, and A.V. with 16 years of clinical neuroradiology experience, respectively), and were iteratively corrected until final approval by one of the neuroradiologists. All manual annotators, including the neuroradiologists, had received prior training and participated in consensus sessions. From the finalized manual segmentations, WT segmentation masks were created by combining all tumor subregions into a single segmentation label. Separately, the ET subregion was extracted. These WT and ET masks were used as ground truth for model training and evaluation.

### Model training and validation

We trained two 3D CNNs using nnU-Net for automated skull-stripping: one with multi-parametric input, and the other with single-parametric input (T1, T1-Gd, T2 or FLAIR). We also trained two different 3D CNNs using nnU-Net for automated brain tumor segmentation: one using only T2 or FLAIR images as input for auto-segmentation of the WT region (without individual tumor subregions), and the other using T1-Gd along with either T2 or FLAIR images for auto-segmentation of the ET region (excluding other tumor subregions). nnU-Net v1 (https://github.com/MIC-DKFZ/nnUNet/tree/nnunetv1) with 5-fold cross validation was trained^32^. Training parameters were: initial learning rate = 0.0, stochastic gradient descent (SGD) with Nesterov momentum (_μ_ = 0.99), and number of epochs = 1000 x 250 minibatches.

A total of 336 subjects were included in the skull-stripping model training, with 153 subjects in the training cohort and a withheld set of 183 subjects in the testing cohort. 489 subjects were included in tumor segmentation model training, with 364 subjects in the training cohort and a withheld set of 125 subjects in the testing cohort. A variety of patient demographics and brain tumor histology were included in this study in order to build robust and generalizable models (Figure 1). The entire image preprocessing, automatic skull-stripping, and tumor segmentation pipeline is presented in Supplemental Figure 1.

### Statistical Analysis

The performances of the different nnU-Net models with respect to the expert manual ground truth segmentations were evaluated using several evaluation metrics, including Dice score (Sørensen-Dice similarity coefficient), sensitivity, and 95% Hausdorff distance.

For skull-stripping, paired t-tests were used to determine any differences in the Dice scores between single- and multi-parametric skull-stripping models, whereas two-sample t-tests were used to compare the differences in Dice scores between different PBT histologies and age-ranges. The correspondence in performance between single- and multi-parametric skull-stripping models was evaluated using Pearson’s correlation.

For tumor segmentation: Paired t-tests were used to compare the performance between T2 or FLAIR inputs for whole tumor segmentation, T1-Gd and T2 or FLAIR inputs for enhancing tumor segmentation for the different histologies. Two-sample t-tests were used for comparing the difference in enhancing tumor volumes for different histologies, using T1-Gd and T2 or T1-Gd and FLAIR inputs. The correlation between ET Dice scores and ET volumes was evaluated using Pearson’s correlation.

## RESULTS

### Skull-stripping Model performance

Supplemental Table 1 shows the resulting Dice scores, sensitivity, and 95% Hausdorff distance for the multi-parametric and single-parametric skull-stripping models for both the whole brain and when selecting only the slices containing sellar/suprasellar regions. For whole brain masks, the multi-parametric and single-parametric models demonstrated similar performance, as indicated by the Dice scores (Supplemental Table 1 and Figure 2A). A similar trend was observed for the sellar/suprasellar slices (Supplemental Table 1 and Figure 2B), with the median Dice scores being slightly higher than those for whole brain masks (Supplemental Table 1). When comparing the performance of the proposed muti-parametric model against the DeepMedic model (which was not trained to include sellar/suprasellar regions) specifically for the sellar/suprasellar slices, the Dice scores (mean±sd (median)) are similar (0.98±0.01 (0.99) vs 0.98±0.01 (0.98)). However, the proposed model showed higher sensitivity (0.98±0.01 (0.99) vs 0.97±0.01 (0.98)) and lower Hausdorff distance (1.06±0.32 (1) vs 1.18±0.36 (1)). The differences in performance between the two models are less pronounced due to the smaller size of the sellar/suprasellar regions compared to the entire brain tissue.

**Figure 2:**
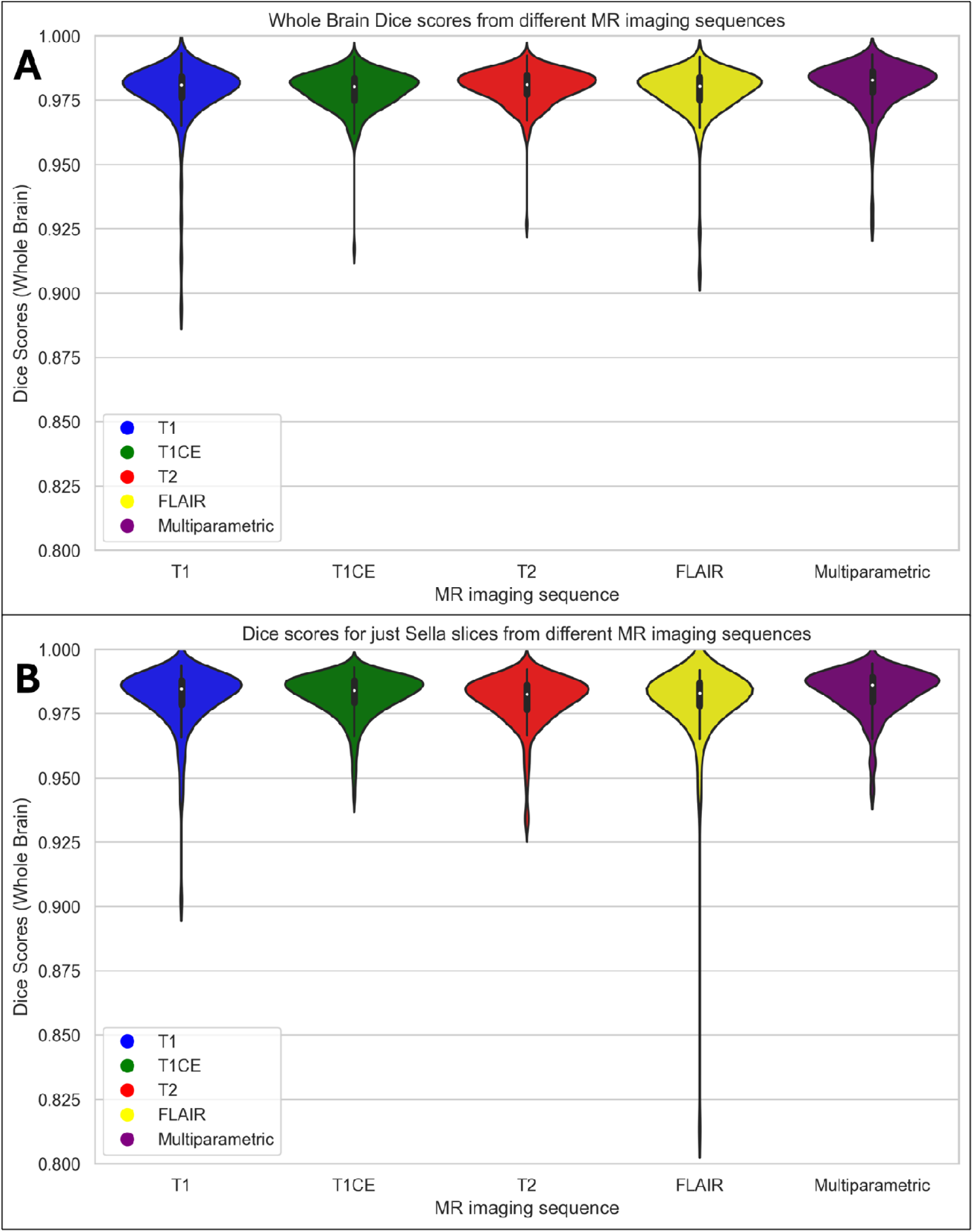
Violin plots showing the distribution of Dice scores for the single-parametric skull-stripping models (T1, T1-Gd, T2 or FLAIR images) compared to the multi-parametric skull-stripping model for whole brain mask (A) and sellar/supraellar slices (B).

The distribution of whole brain Dice scores using the multi-parametric and single-parametric skull-stripping models for different brain tumor histologies – low-grade glioma (LGG), medulloblastoma, diffuse midline glioma (DMG), and other histologies including high-grade glioma – astrocytoma (HGG), ependymoma, ganglioglioma – shows that both models performed similarly well (Supplemental Figure 2). Performance was largely affected by one DMG subject in the single-parametric model with T1 input, which had a whole brain Dice score of 0.89, leading to a slightly less dense group-level distribution.

Comparison of the Dice scores between the multi-parametric and single-parametric models for whole brain and sellar slices showed significant correlation (p<0.05), demonstrating similar performance for skull-stripping in both whole brain and sellar slices (Supplemental Figures 3 and 4).

Whole brain Dice scores of the the multi-parametric model showed no significant difference across different age ranges (0 – 3, 3 – 13, and 13 – 18 years) (all p>0.05), suggesting the model’s generalizability across all pediatric age groups (Supplemental Figure 5).

Supplemental Figure 6 depicts representative pre-processed brain MR images overlaid with ground truth brain masks along with predicted whole brain masks from single-parametric and multi-parametric skull stripping models. The successful performance of multi-parametric model on craniopharyngioma and germinoma, which originate from the sellar/suprasellar regions, is also shown.

We further tested the impact of our proposed skull-stripping model on automated tumor segmentation by comparing skull-stripped images to non-skull-stripped images in data from 12 subjects with sellar/suprasellar tumors. Supplemental Table 2 shows the mean ± sd (median) values for Dice score, sensitivity, and 95% Hausdorff distance metrics for WT segmentation in both skull-stripped and non-skull-stripped images. The results indicate no significant difference, based on the Wilcoxon signed-rank test (p=0.18), suggesting no adverse impact of skull-stripping on a downstream task. Finally, qualitive comparison of the nnU-Net-based skull-stripping model with previously reported DeepMedic-based skull-stripping model, reveals that the nnU-Net-based skull-stripping model performs better at including sellar/suprasellar areas, frontal lobe, and brainstem from the brain tumor as part of the extracted brain tissue region (Figure 3).

**Figure 3:**
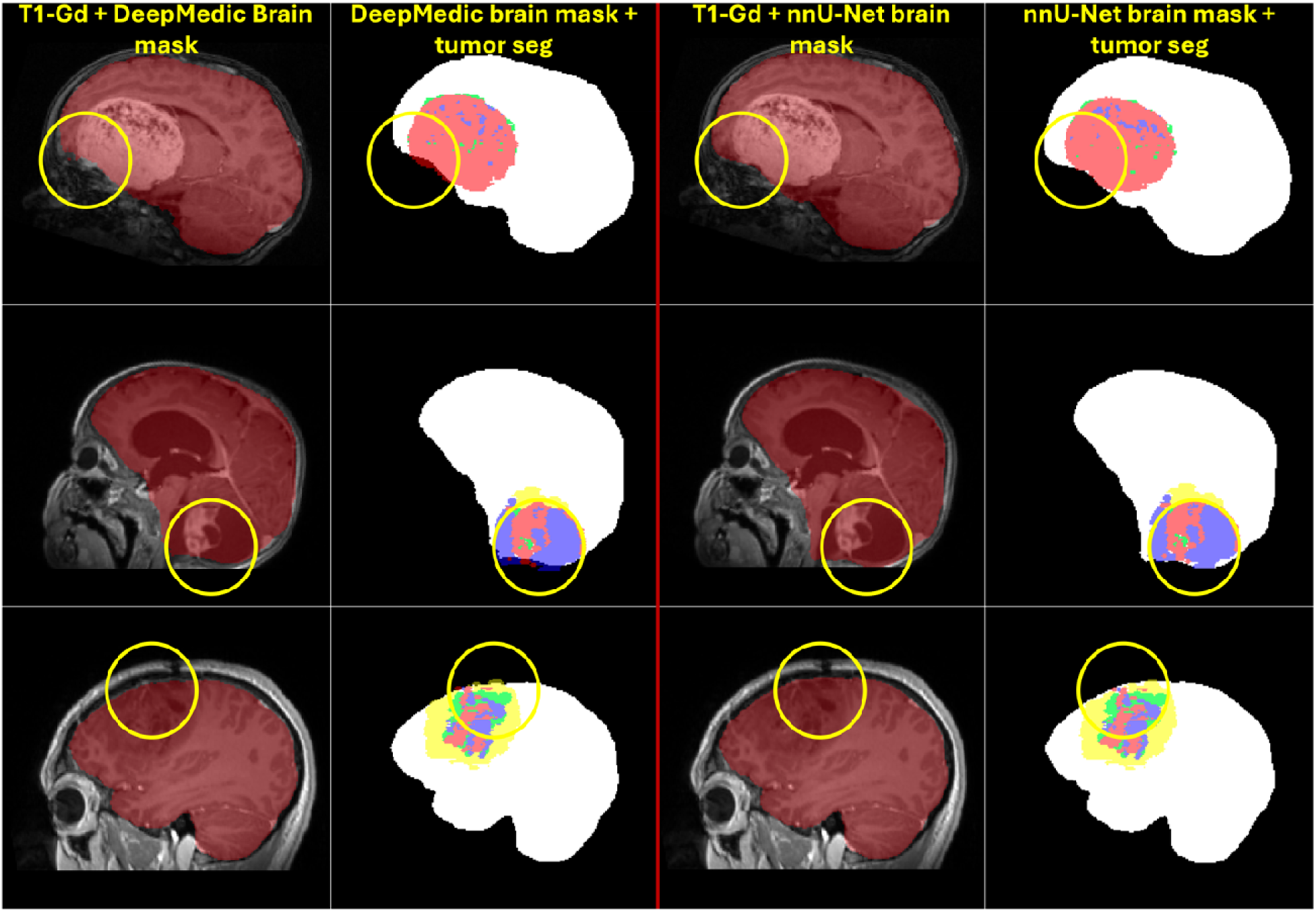
Examples of skull-stripping performance using the multi-parametric nnU-Net-based model compared to an earlier pediatric DeepMedic-based skull-stripping model. The nnU-Net-based model shows improved performance, successfully segmenting tumor regions that the DeepMedic-based model fails to include as part of the brain tissue.

### Tumor segmentation Model performance

Table 1 shows the Dice scores, sensitivity and 95% Hausdorff distance for 1) T2 or FLAIR model – WT region and 2) T1-Gd and either T2 or FLAIR model – ET region. The distribution of Dice scores from the T2 and FLAIR models showed no significant difference between various tumor histologies for both T2 and FLAIR images (Figure 4A). An example of preprocessed MRI sequences, images overlaid with the ground truth segmentation, and predicted segmentations for the T1-Gd and FLAIR model, T1-Gd and T2 model, and T2 or FLAIR model is shown (Figure 5 A,B and C, respectively).

**Figure 4:**
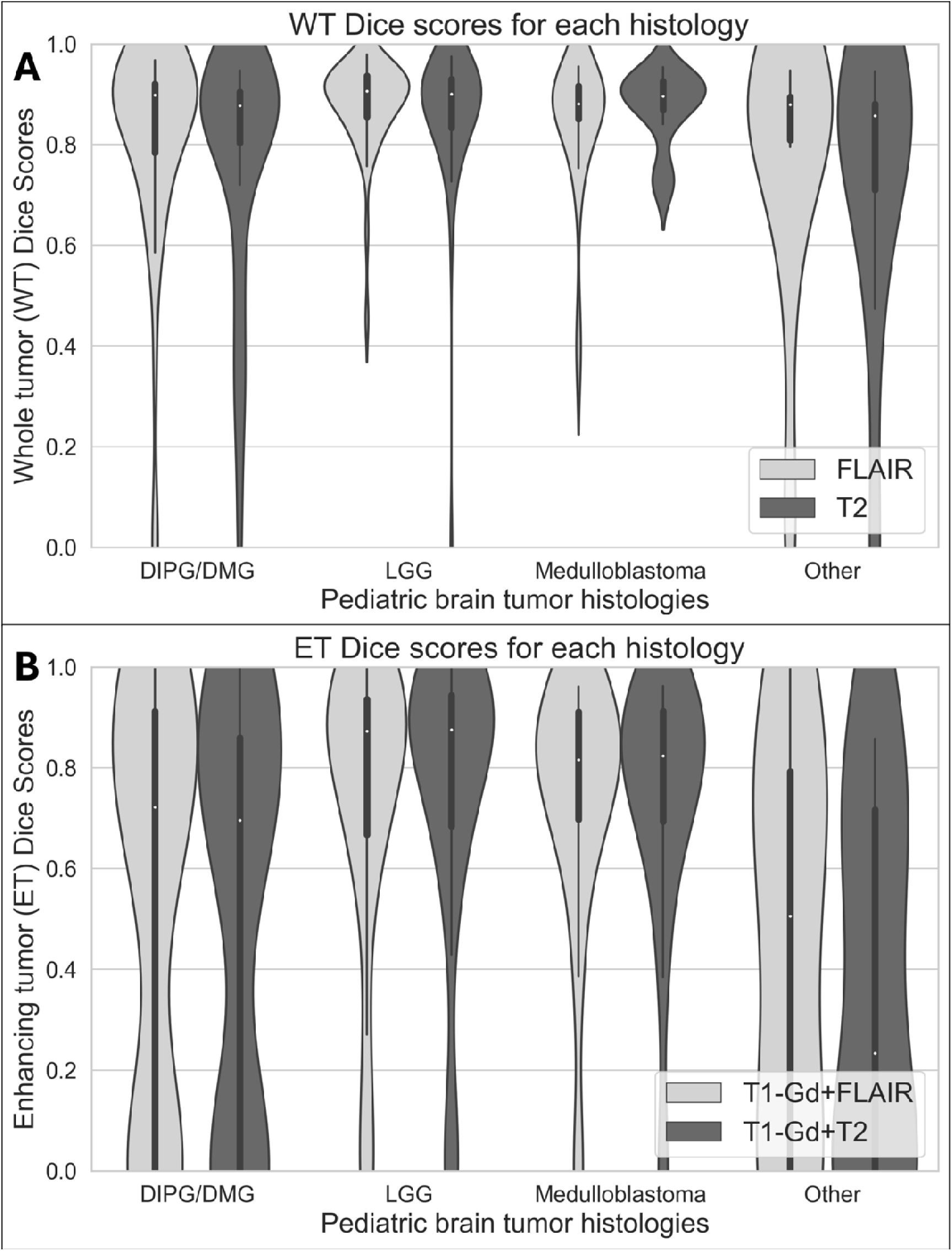
Violin plots showing the distribution of whole tumor (WT) Dice scores using the T2 or FLAIR tumor segmentation model for different histologies (A). There is no significant difference in the Dice scores between T2 and FLAIR inputs for different tumor histologies. Violin plots showing the distribution of enhancing tumor (ET) Dice scores using the T1-Gd and T2 or FLAIR tumor segmentation models for the different histologies (B). ET Dice scores from LGG and medulloblstoma patients were siginificantly higher than those from DIPG/DMG and Other histologies (all p<0.0.5).

**Figure 5:**
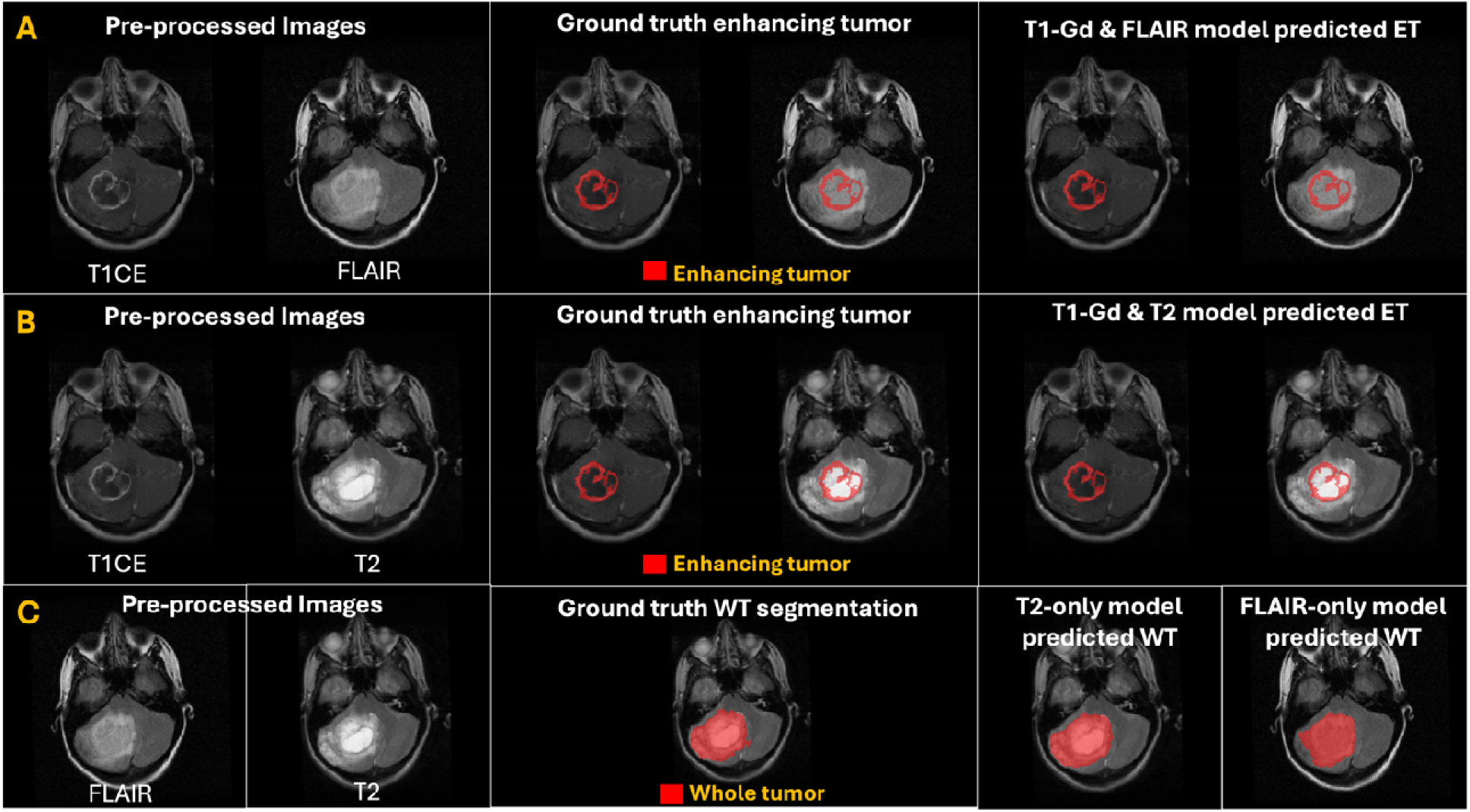
(A) and (B) show example brain MR images overlaid with ground truth segmentation labels and model-predicted enhancing tumor segmentation labels for T1-Gd and FLAIR, and T1-Gd and T2, respectively. (C) shows results for the T2 or FLAIR model for segmenting WT.

**Table 1:** Performance metrics for T2 or FLAIR whole tumor segmentation models, and T1-Gd and T2 or FLAIR enhancing tumor segmentation models.

| Model & Region /<br>Metric | Dice score<br>Mean±sd (Median) | Sensitivity<br>Mean±sd (Median) | 95% Hausdorff<br>distance<br>Mean ± sd (Median) |
| --- | --- | --- | --- |
| **One-sequence models – WT** |  |  |  |
| FLAIR-only | 0.84±0.17 (0.90) | 0.83±0.18 (0.88) | 8.09±13.20 (3.32) |
| T2-only | 0.82±0.19 (0.89) | 0.8±0.21 (0.88) | 8.24±12.79 (3.61) |
| **Two-sequence models – ET** |  |  |  |
| T1-Gd and FLAIR | 0.65±0.35 (0.79) | 0.76±0.26 (0.86) | 6.41±9.25 (3) |
| T1-Gd and T2 | 0.64±0.36 (0.79) | 0.76±0.26 (0.85) | 6.09±9.33 (2.83) |

The distribution of ET Dice scores using the T1-Gd with T2 or FLAIR inputs for different PBT histologies showed that ET Dice scores for DMG are significantly lower than those for LGG and medulloblastoma (Figure 4B). Two sample t-tests indicated significant differences in ET Dice scores between DMG and LGG (p=0.004 for T1-Gd and T2, p=0.018 for T1-Gd and FLAIR), between DMG and medulloblastoma (p=0.02 for T1-Gd and T2, p=0.037 for T1-Gd and FLAIR). Additionally, significant differences were found between tumors categorized as “Other Histologies” and LGG (p=0.0003 for T1-Gd and T2, p=0.011 for T1-Gd and FLAIR) and “Other Histologies” and medulloblastoma (p=0.0008 for T1-Gd and T2, p=0.011 for T1-Gd and FLAIR). LGG and medulloblastoma subjects had significantly higher ET volumes than DMG and “Other Histologies”, indicating the model’s superior performance in histologies where the ET region is more prevalent and tumor size is larger (Supplemental Figures 7A and 7B). A scatter plot comparing ET Dice scores with ground truth ET volumes showed a low but significant correlation (r=0.33, p<0.05) (Supplemental Figure 7C).

## DISCUSSION

In this study, we aimed to develop generalizable, pediatric-specific automated methods for skull-stripping that includes the sellar/suprasellar regions and an automated tumor segmentation approach for use in limited data contexts. Our study demonstrated excellent results for skull-stripping for both the multi-parametric and single-parametric models, with similar performance on the whole brain mask and sellar/suprasellar slices. The brain tumor segmentation models presented in this study build on our previous work, enabling the evaluation of imaging sessions of PBTs without multi-parametric MRI acquisitions. Additionally, these models allow for the segmentation of the non-enhancing component by subtracting ET from WT in cases where T1-Gd and either T2 or FLAIR images are available.

This study utilized the most comprehensive pediatric brain MRI dataset for training skull-stripping and tumor auto-segmentation models to date, based on the diversity of tumor histologies included in both training and testing. The dataset was multi-institutional, encompassing a wide range of MRI scanner field strengths and manufacturers (see Supplemental Table 3). It included a variety of brain tumor types, with the major ones being LGG, medulloblastomas, and DMG, as well as other types such as HGG, ependymomas, germinomas, craniopharyngiomas, and other rare tumors.

Our automated tumor segmentation models using only T2 or FLAIR sequences achieved high accuracy in WT region segmentation (median Dice scores: 0.9 for FLAIR-only input and 0.89 for T2-only input). The results for ET region segmentation using combinations of T1-Gd with T2 or FLAIR sequences were more moderate (median Dice scores: 0.79 for both T1-Gd & T2 and T1-Gd & FLAIR models).

Comparing our results to the BraTS-PEDs 2023 challenge ^36^, our WT and ET segmentation models performed better than the top-performing models. For WT segmentation using our T2 or FLAIR models compared to the top-performing BraTS-PEDS 2023 teams, Dice score was 0.84±0.17 (0.90) vs. 0.84±0.16 (0.87), sensitivity was 0.83±0.18 (0.88) vs. 0.8±0.09 (0.82), and 95% Hausdorff distance was 8.09±13.20 (3.32) vs. 18.05±62.77 (4.30). Similarly, for ET segmentation using our T1-Gd with T2 or FLAIR models compared to the top-performing BraTS-PEDS 2023 teams, Dice score was 0.65±0.35 (0.79) vs. 0.65±0.32 (0.74), sensitivity was 0.76±0.26 (0.86) vs. 0.7±0.18 (0.74) and 95% Hausdorff distance was 6.41±9.25 (3) vs. 43.89±108.59 (3.67). These results collectively highlight that while our models used limited imaging sets for segmentation of WT and ET regions, as opposed to the methods submitted to the BraTS-PEDs 2023 that use multi-parametric MRI, they achieve better performance.

While numerous studies have focused on skull-stripping in adult brain MRI scans, only a few have addressed this task using pediatric brain MRI scans. Kazerooni *et al.* demonstrated Dice scores of 0.98 using the DeepMedic architecture for skull-stripping with multi-parametric MR images, which is comparable to our results. However, their study used a smaller dataset of 21 cases for inference, did not include single-parametric models, and did not encompass the sellar/suprasellar regions in the brain masks ^16^. Kim *et al.* reported whole brain Dice scores in the range of 0.79-0.8 using VUNO Med-DeepBrain for subjects with SCN1A mutations (n=21) and healthy subjects (n=42) in a multi-institutional and multi-scanner dataset. However, their study did not include any subjects with brain tumors and used a significantly smaller dataset compared to ours ^19^. Chen et al. used ANUBEX based on nnU-Net for skull-stripping in neonates and compared it against five other deep learning models, achieving Dice scores in the range of 0.92 to 0.96 ^42^. However, their model only used T1 images as input and was tested on a small withheld dataset of 39 subjects. In contrast, our study demonstrated excellent skull-stripping results using both multi-parametric and single-parametric models, outperforming previously published studies. Additionally, our study showed accurate skull-stripping performance across different pediatric age groups (0-3, 3-13, and 13-18 years), highlighting its robustness to the structural and signal intensity changes in the skull due to child development.

While Dice scores for skull-stripping with sellar/suprasellar region inclusion do not directly inform on the performance of downstream analyses, a comparison between the DeepMedic skull-stripping model (not trained to include sellar/suprasellar regions) and our proposed nnU-Net-based model indicates that the latter is qualitatively better at including brain tumor regions from the sellar/suprasellar areas, frontal lobe, and brainstem as part of the extracted brain tissue. Furthermore, using either skull-stripped images or non skull-stripped images as input to the automated tumor segmentation model did not impact its performance for tumors located in the sellar/suprasellar regions.

A few studies in the literature have investigated the use of T2 or FLAIR images for WT segmentation in pediatric populations. Boyd et al. trained and compared multiple stepwise transfer learning models based on nnU-Net for WT segmentation using T2 images ^43^. The best performing transfer-encoder model had a median Dice score of 0.877 for the internal validation set, whereas the external validation set had a median Dice score of 0.833. These median Dice scores are lower compared to the median WT Dice score of 0.89 using T2 inputs for the tumor segmentation model presented in our study. Additionally, the transfer-encoder stepwise transfer learning model was only trained on LGG cases, whereas our multi-institutional dataset was larger and trained on a wider range of PBT histologies. Furthermore, Vafaeikia et al. trained a 2 step U-Net based deep learning model for WT segmentation using just FLAIR images ^44^. They reported a mean Dice score of 0.795, which is lower than the mean Dice score of 0.84 reported in our study for WT segmentation using just FLAIR images. The 2 step U-Net model was trained only on LGG patients and included data from a single institution, compared to the wider range of PBT histologies and multi-institutional dataset included in the present study. To the best of our knowledge, there is only one study in literature that explored the use of one or two input MRI sequences for segmenting the enhancing tumor region in case of PBT. Peng et al. used T1-Gd and T2 images to train a U-Net model for ET segmentation from 638 pre-operative PBT patients^45^. They reported a mean and median Dice scores of 0.724 and 0.843, respectively, compared to our proposed model, which, for the same inputs had resulted in a mean and median Dice scores of 0.64 and 0.79 respectively. While the U-Net model was trained on larger dataset and demonstrated better results, it is important to note that our proposed model works with T1-Gd and T2 or FLAIR inputs

Our study had a few limitations that are important to note. In subjects with smaller enhancing tumor subregion volumes, even slight inaccuracies in model prediction can push the Dice scores to extreme values (close or equal to 0) due to the low number of voxels being compared. This disproportionately penalizes model performance and biases subsequent statistical analysis. Moreover, weak enchancement and poor-quality T1-Gd scans can complicate the segmentation of the ET region ^10^. Despite these limitations, we believe that the proposed ET segmentation model will help reduce the burden of manual segmentation by providing a reliable initial prediction of the ET region, which can then be reviewed and modified, by experienced radiologists.

Future work will include training a separate tumor segmentation model for post-operative subjects, perhaps with an additional label for the resected tumor. Incorporating intraorbital tumors and fine-tuning the model with data from specific histologies, such as DMG, will improve its performance in segmenting tumor sub-regions, beneficial for many applications. Additionally, we plan to apply our tumor segmentation models to clinical trial studies that monitor tumor response to treatment, to further demonstrate their generalizability.

## CONCLUSION

In summary, this study presents enhanced skull-stripping and tumor segmentation models that are more generalizable across various PBT histologies and adaptable to limited MRI sequence availability. The proposed skull-stripping models can support applications such as synthesizing missing MRI sequences using generative adversarial networks ^46^ and extracting radiomic features, both of which depend on accurate and comprehensive brain tissue segmentation, including the entirety of tumor.

The single-parametric skull-stripping models, as well as one-input (T2 or FLAIR) whole tumor segmentation and two-input (T1-Gd and T2 or FLAIR) enhancing tumor segmentation, enable the inclusion of cases with incomplete multi-parametric image sets in limited data context. These advancements facilitate more extensive clinical translation and improved assessment of PBTs.

## Data Availability

All data produced in the present study are available upon reasonable request to the authors. All image processing tools that were used in this study are freely available for public use (CaPTk, https://www.cbica.upenn.edu/captk; ITK-SNAP, https://www.itksnap.org). The pediatric pre-processing and segmentation pipeline, along with pre-trained nnU-Net skull-stripping and tumor segmentation models are publicly available online at [https://github.com/d3b-center/peds-brain-seg-pipeline-public]. The stand-alone skull-stripping models are publicly available at [https://github.com/d3b-center/peds-brain-auto-skull-strip].

https://github.com/d3b-center/peds-brain-auto-skull-strip

## Abbreviations

CNS = Central nervous system; PBT = Pediatric brain tumors; CNN = Convolutional Neural Networks; CBTN = Children’s Brain Tumor Network; WT = whole tumor; ET = enhancing tumor; NET = non-enhancing tumor; CC = cystic component; ED = edema; T1-Gd = Contrast-enhanced T1-weighted image; FLAIR = fluid attenuated inversion recovery.

## AUTHORSHIP

Conceptualization: A.F.K., A.N., A.V., Resources: P.B.S., A.C.R., A.N., and A.F.K., Data curation: D.B.G., N.K., A.M.F., A.G., N.K., W.T., S.H., H.A., and K.V., Writing – original draft preparation: D.B.G., N.K., A.F.K., Writing – review and editing: A.N., J.B.W., A.M.F., A.F.K., D.B.G., N.K., and A.V., Supervision: A.F.K., A.N., Funding acquisition: A.F.K., P.B.S. and A.C.R.

## DATA AND CODE AVAILABILITY

All image processing tools that were used in this study are freely available for public use (CaPTk, https://www.cbica.upenn.edu/captk; ITK-SNAP, https://www.itksnap.org). The pediatric pre-processing and segmentation pipeline, along with pre-trained nnU-Net skull-stripping and tumor segmentation models are publicly available online at [https://github.com/d3b-center/peds-brain-seg-pipeline-public]. The stand-alone skull-stripping models are publicly available at [https://github.com/d3b-center/peds-brain-auto-skull-strip]. Additionally, the data used for training/testing the models can be made available by the corresponding author upon reasonable request

## REFERENCES

1. Hossain, M.J., Xiao, W., Tayeb, M. & Khan, S. Epidemiology and prognostic factors of pediatric brain tumor survival in the US: Evidence from four decades of population data. Cancer epidemiology 72(2021/06).

2. Racial and ethnic differences in survival of pediatric patients with brain and central nervous system cancer in the United States - PubMed. Pediatric blood & cancer 66(2019 Feb).

3. Claus, E.B. & Black, P.M. Survival rates and patterns of care for patients diagnosed with supratentorial low-grade gliomas. Cancer 106(2006/03/15).

4. Global survival trends for brain tumors, by histology: Analysis of individual records for 67,776 children diagnosed in 61 countries during 2000-2014 (CONCORD-3) - PubMed. Neuro-oncology 25(03/14/2023).

5. Ostrom, Q.T., et al. CBTRUS Statistical Report: Pediatric Brain Tumor Foundation Childhood and Adolescent Primary Brain and Other Central Nervous System Tumors Diagnosed in the United States in 2014–2018. Neuro-Oncology 24, iii1-iii38 (2022).

6. Pediatric Brain Tumors - PubMed. Neurologic clinics 36(2018 Aug).

7. Frühwald, M.C. & Rutkowski, S. Tumors of the Central Nervous System in Children and Adolescents. Deutsches Ärzteblatt International 108(2011/06).

8. The 2021 WHO Classification of Tumors of the Central Nervous System: a summary - PubMed. Neuro-oncology 23(08/02/2021).

9. Simarro, J., et al. A deep learning model for brain segmentation across pediatric and adult populations. Scientific Reports 14, 11735 (2024).

10. Familiar, A.M., et al. Towards consistency in pediatric brain tumor measurements: Challenges, solutions, and the role of artificial intelligence-based segmentation. Neuro-Oncology (2024).

11. Veiga-Canuto, D., et al. Comparative Multicentric Evaluation of Inter-Observer Variability in Manual and Automatic Segmentation of Neuroblastic Tumors in Magnetic Resonance Images. Cancers 14, 3648 (2022).

12. Khalighi, S., et al. Artificial intelligence in neuro-oncology: advances and challenges in brain tumor diagnosis, prognosis, and precision treatment. npj Precision Oncology 8, 80 (2024).

13. Liu, Z., et al. Deep learning based brain tumor segmentation: a survey. Complex & Intelligent Systems 2022 9:1 92022-07-09).

14. Boyd, A., et al. Expert-level pediatric brain tumor segmentation in a limited data scenario with stepwise transfer learning. medRxiv (2023).

15. Vossough, A., et al. Training and Comparison of nnU-Net and DeepMedic Methods for Autosegmentation of Pediatric Brain Tumors. American Journal of Neuroradiology (2024-05-09).

16. Fathi Kazerooni, A., et al. Automated tumor segmentation and brain tissue extraction from multiparametric MRI of pediatric brain tumors: A multi-institutional study. Neuro-Oncology Advances 5(2023/01/01).

17. Liu, X., et al. From adult to pediatric: deep learning-based automatic segmentation of rare pediatric brain tumors. 10.1117/12.265424512465(2023/04/07).

18. Aggarwal, M., Tiwari, A.K., Sarathi, M.P. & Bijalwan, A. An early detection and segmentation of Brain Tumor using Deep Neural Network. BMC Medical Informatics and Decision Making 23(2023).

19. Kim, M.-J., et al. Deep learning-based, fully automated, pediatric brain segmentation. Scientific Reports 2024 14:1 14(2024-02-22).

20. Hoopes, A., Mora, J.S., Dalca, A.V., Fischl, B. & Hoffmann, M. SynthStrip: skull-stripping for any brain image. NeuroImage 260, 119474 (2022).

21. Kalavathi, P. & Prasath, V.B. Methods on Skull Stripping of MRI Head Scan Images-a Review. J Digit Imaging 29, 365–379 (2016).

22. Fatima, A., Shahid, A.R., Raza, B., Madni, T.M. & Janjua, U.I. State-of-the-Art Traditional to the Machine- and Deep-Learning-Based Skull Stripping Techniques, Models, and Algorithms. J Digit Imaging 33, 1443–1464 (2020).

23. Hwang, H., Rehman, H.Z.U. & Lee, S. 3D U-Net for Skull Stripping in Brain MRI. Applied Sciences 9, 569 (2019).

24. Maia, R., et al. Neuroimaging of pediatric tumors of the sellar region—A review in light of the 2021 WHO classification of tumors of the central nervous system. Frontiers in Pediatrics 11(2023).

25. Al-Bader, D., Hasan, A. & Behbehani, R. Sellar masses: diagnosis and treatment. Frontiers in Ophthalmology 2(2022).

26. Familiar, A.M., et al. A multi-institutional pediatric dataset of clinical radiology MRIs by the Children’s Brain Tumor Network. ArXiv (2023).

27. Gilligan, L.A., DeWire-Schottmiller, M.D., Fouladi, M., DeBlank, P. & Leach, J.L. Tumor Response Assessment in Diffuse Intrinsic Pontine Glioma: Comparison of Semiautomated Volumetric, Semiautomated Linear, and Manual Linear Tumor Measurement Strategies. AJNR Am J Neuroradiol 41, 866–873 (2020).

28. Fangusaro, J., et al. Response assessment in paediatric low-grade glioma: recommendations from the Response Assessment in Pediatric Neuro-Oncology (RAPNO) working group. The Lancet Oncology 21, e305–e316 (2020).

29. Erker, C., et al. Response assessment in paediatric high-grade glioma: recommendations from the Response Assessment in Pediatric Neuro-Oncology (RAPNO) working group. The Lancet Oncology 21, e317–e329 (2020).

30. Lee, W.-K., et al. Lesion delineation framework for vestibular schwannoma, meningioma and brain metastasis for gamma knife radiosurgery using stereotactic magnetic resonance images. Computer Methods and Programs in Biomedicine 229, 107311 (2023).

31. Lee, W.-K., et al. Combining analysis of multi-parametric MR images into a convolutional neural network: Precise target delineation for vestibular schwannoma treatment planning. Artificial Intelligence in Medicine 107, 101911 (2020).

32. Isensee, F., et al. nnU-Net: a self-configuring method for deep learning-based biomedical image segmentation. Nature Methods 2020 18:2 18(2020-12-07).

33. Brain tumor segmentation with advanced nnU-Net: Pediatrics and adults tumors. Neuroscience Informatics 4(2024/06/01).

34. Boer, M.d., et al. NnU-Net versus mesh growing algorithm as a tool for the robust and timely segmentation of neurosurgical 3D images in contrast-enhanced T1 MRI scans. Acta Neurochirurgica 166(2024).

35. Lilly, J.V., et al. The children’s brain tumor network (CBTN) - Accelerating research in pediatric central nervous system tumors through collaboration and open science. Neoplasia (New York, N.Y.) 35(2023/01).

36. Kazerooni, A.F., et al. BraTS-PEDs: Results of the Multi-Consortium International Pediatric Brain Tumor Segmentation Challenge 2023. arXiv preprint arXiv:2407.08855 (2024).

37. The SRI24 multichannel atlas of normal adult human brain structure - PubMed. Human brain mapping 31(2010 May).

38. Cancer imaging phenomics toolkit: quantitative imaging analytics for precision diagnostics and predictive modeling of clinical outcome - PubMed. Journal of medical imaging (Bellingham, Wash.) 5(2018 Jan).

39. Pati, S., et al. The Cancer Imaging Phenomics Toolkit (CaPTk): Technical Overview. Brainlesion : glioma, multiple sclerosis, stroke and traumatic brain injuries. BrainLes (Workshop) 11993(2020).

40. Thakur, S.P., et al. Skull-Stripping of Glioblastoma MRI Scans Using 3D Deep Learning. Brainlesion : glioma, multiple sclerosis, stroke and traumatic brain injuries. BrainLes (Workshop) 11992(2019/10).

41. Yushkevich, P.A., Gao, Y. & Gerig, G. ITK-SNAP: an interactive tool for semi-automatic segmentation of multi-modality biomedical images. Conference proceedings : … Annual International Conference of the IEEE Engineering in Medicine and Biology Society. IEEE Engineering in Medicine and Biology Society. Annual Conference 2016(2016/08).

42. Chen, J.V., et al. Automated neonatal nnU-Net brain MRI extractor trained on a large multi-institutional dataset. Scientific Reports 2024 14:1 14(2024-02-26).

43. Boyd, A., et al. Expert-level pediatric brain tumor segmentation in a limited data scenario with stepwise transfer learning. medRxiv (2023).

44. Vafaeikia, P., et al. MRI-Based End-To-End Pediatric Low-Grade Glioma Segmentation and Classification. Canadian Association of Radiologists Journal 75, 153–160 (2024).

45. Peng, J., et al. Deep learning-based automatic tumor burden assessment of pediatric high-grade gliomas, medulloblastomas, and other leptomeningeal seeding tumors. Neuro-Oncology 24, 289–299 (2021).

46. Chrysochoou, D., et al. IMG-08. SYNTHESIZING MISSING MRI SEQUENCES IN PEDIATRIC BRAIN TUMORS USING GENERATIVE ADVERSARIAL NETWORKS; TOWARDS IMPROVED VOLUMETRIC TUMOR ASSESSMENT. Neuro-Oncology 26, 0–0 (2024).

