## Supplemental for "Automated Pediatric Brain Tumor Imaging Assessment Tool from CBTN: Enhancing Suprasellar Region Inclusion and Managing Limited Data with Deep Learning"

**SUPPLEMENTAL FIGURES:**

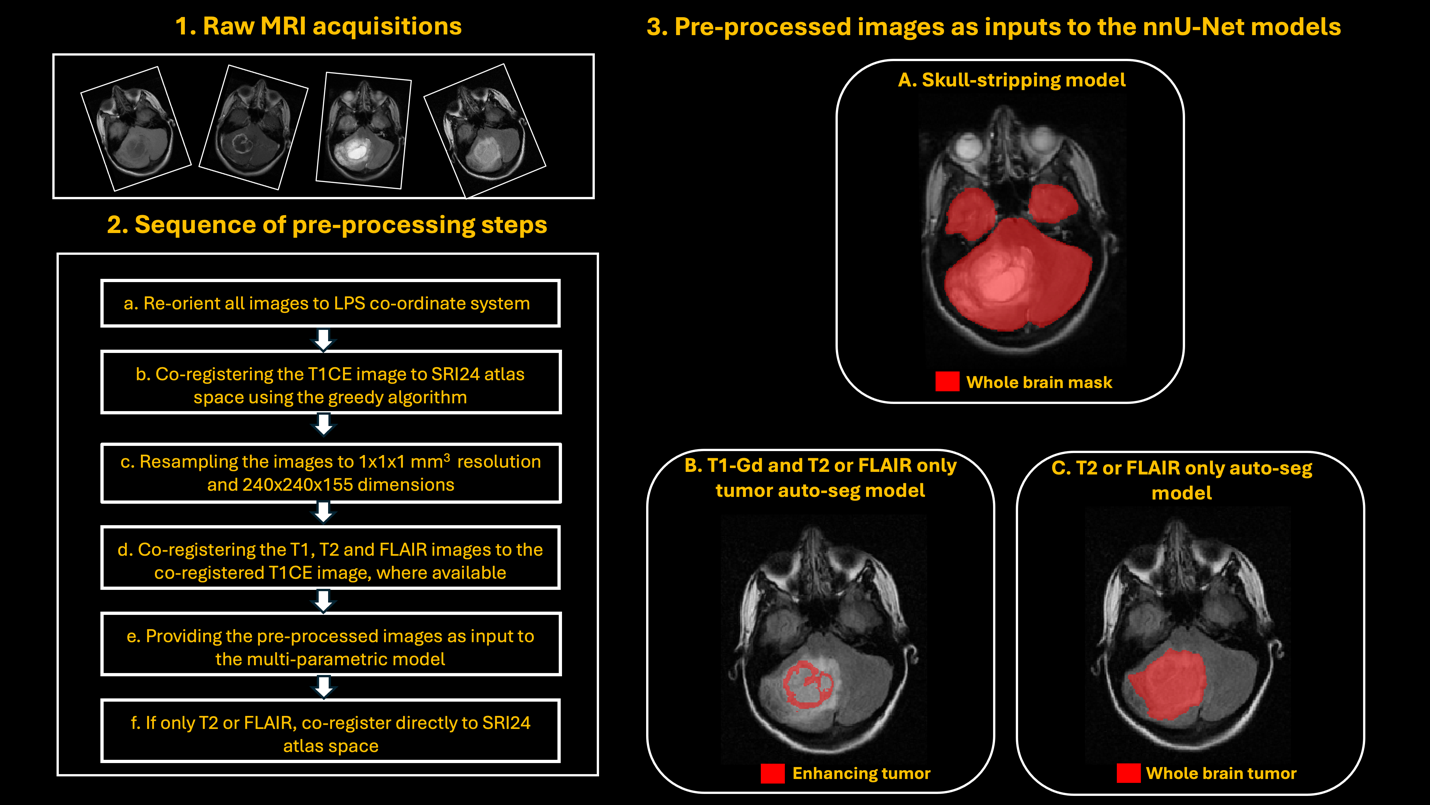

**Supplemental Figure 1.** Workflow of the entire end-to-end pediatric brain MRI pre-processing and fully automatic skull-stripping and tumor segmentation models. 1) Input raw brain MRI acquisitions, 2) Sequential steps for pre-processing the raw input images, along with the resulting pre-processed images, and 3) Pre-processed images as inputs to various nnU-Net models: A) Automatic skull-stripping, B) T1-Gd and T2 or FLAIR models for segmenting enhancing tumor (ET) and C) T2 or FLAIR model for segmenting just the whole tumor (WT) region.

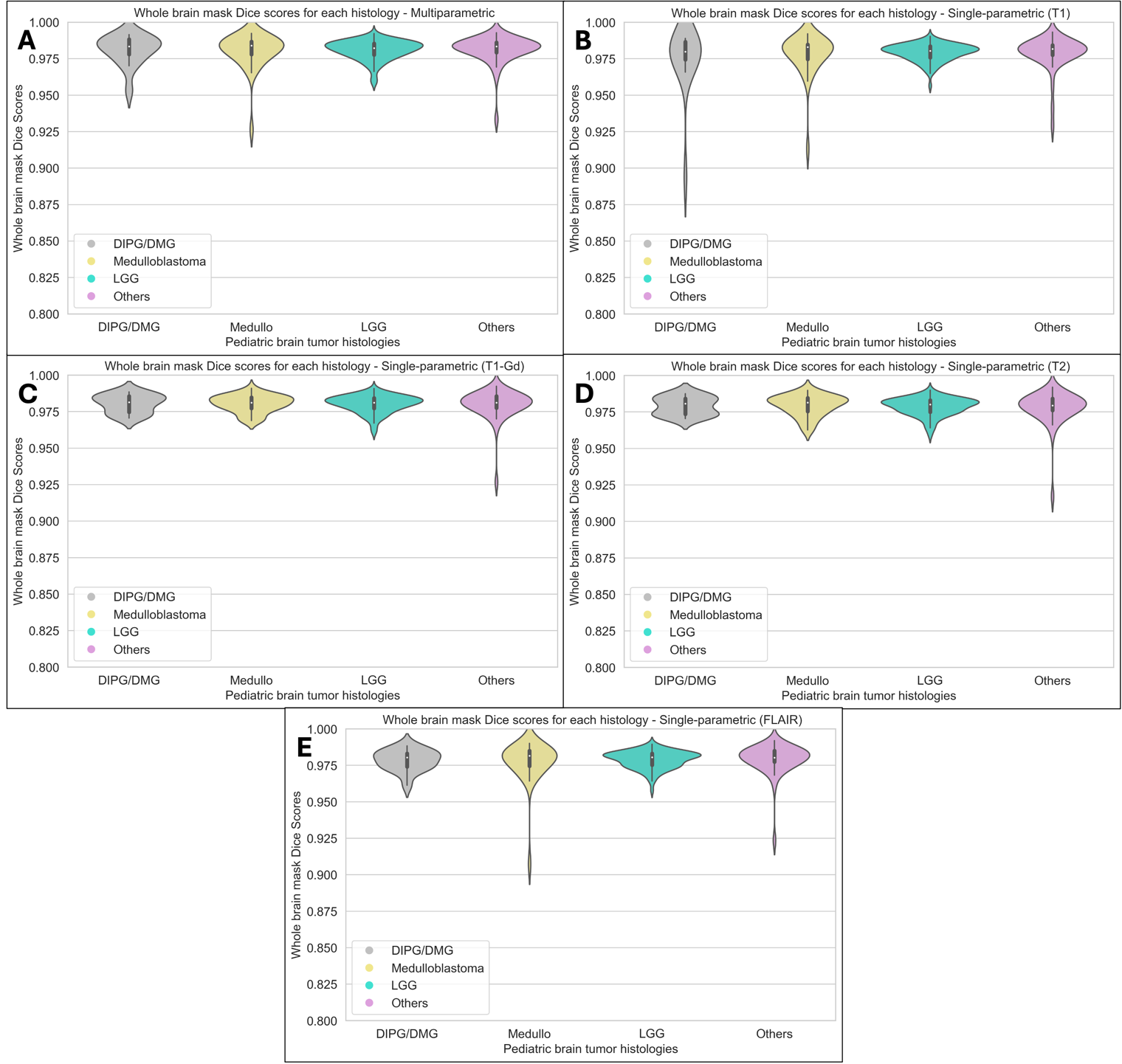

**Supplemental Figure 2.** Violin plots showing the distribution of whole brain mask Dice scores obtained using the multi-parametric skull-stripping model (A), and single-parametric skull-stripping model for each modality (B – T1, C – T1-Gd, D – T2 and E – FLAIR). These plots compare performance among various brain tumor histologies: DMG, medulloblastoma, LGG and Other histologies.

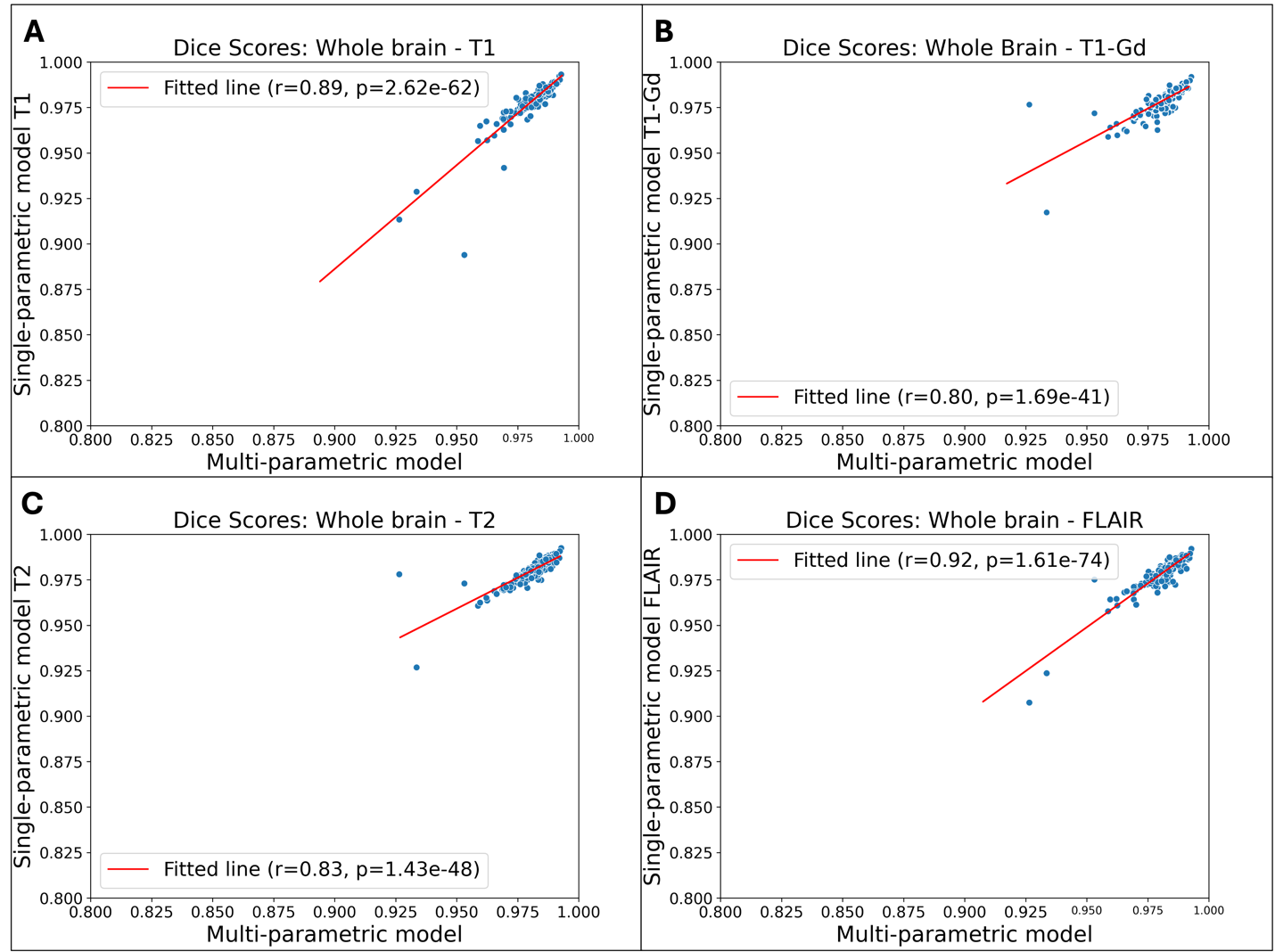

**Supplemental Figure 3.** Scatter plots showing the correlation between Dice scores obtained from the multi-parametric and single-parametric skull-stripping models with individual MRI sequences as inputs. There is a significant correlation between whole brain Dice scores obtained from the multi-parametric and each single-parametric model.

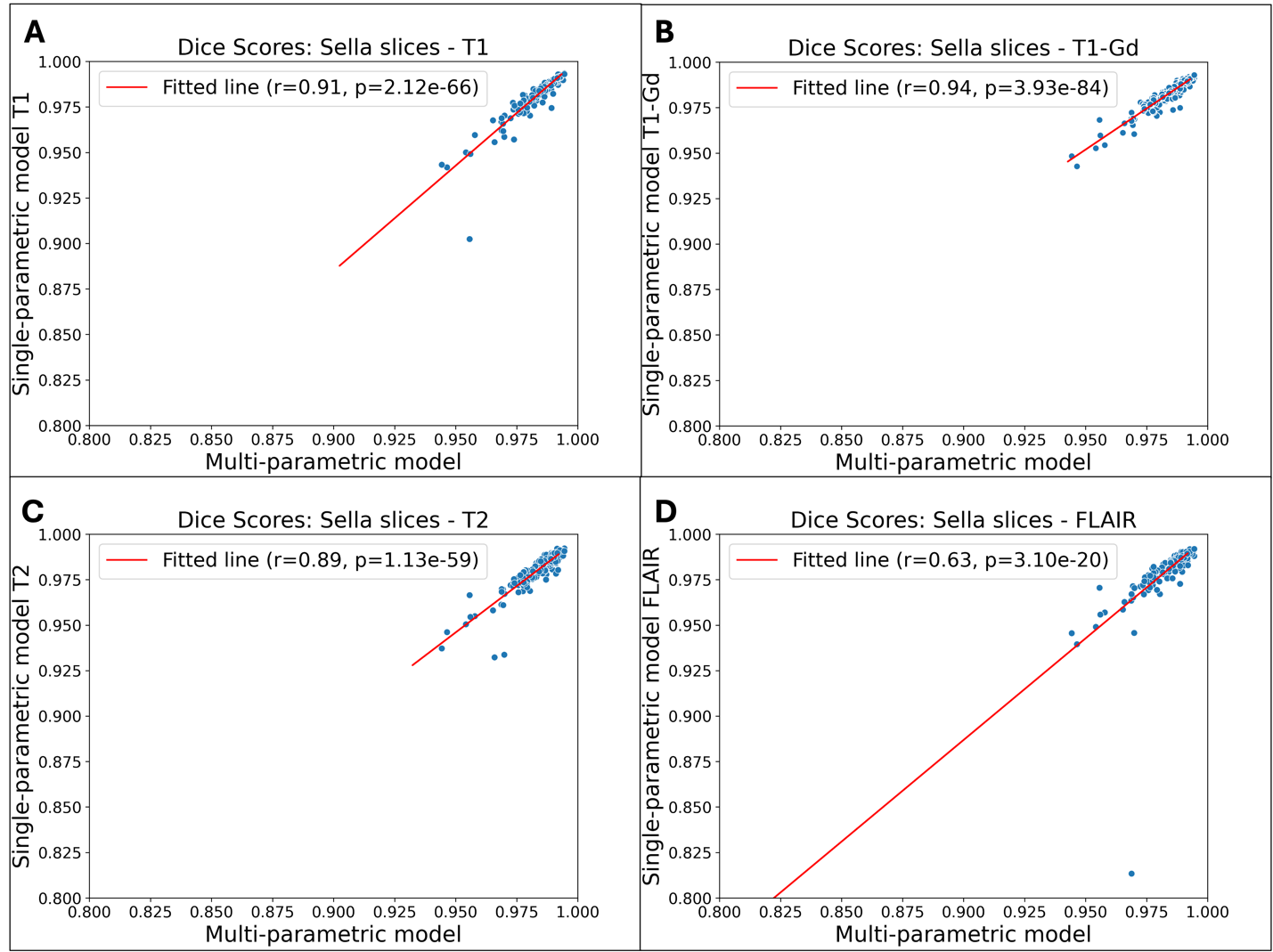

**Supplemental Figure 4.** Scatter plots showing the correlation between Dice scores obtained from the multi-parametric model and single-parametric models with individual MRI sequences as input for the sellar slices. There is a significant correlation between Dice scores obtained from the multi-parametric and each single-parametric model in the sellar/suprasellar regions.

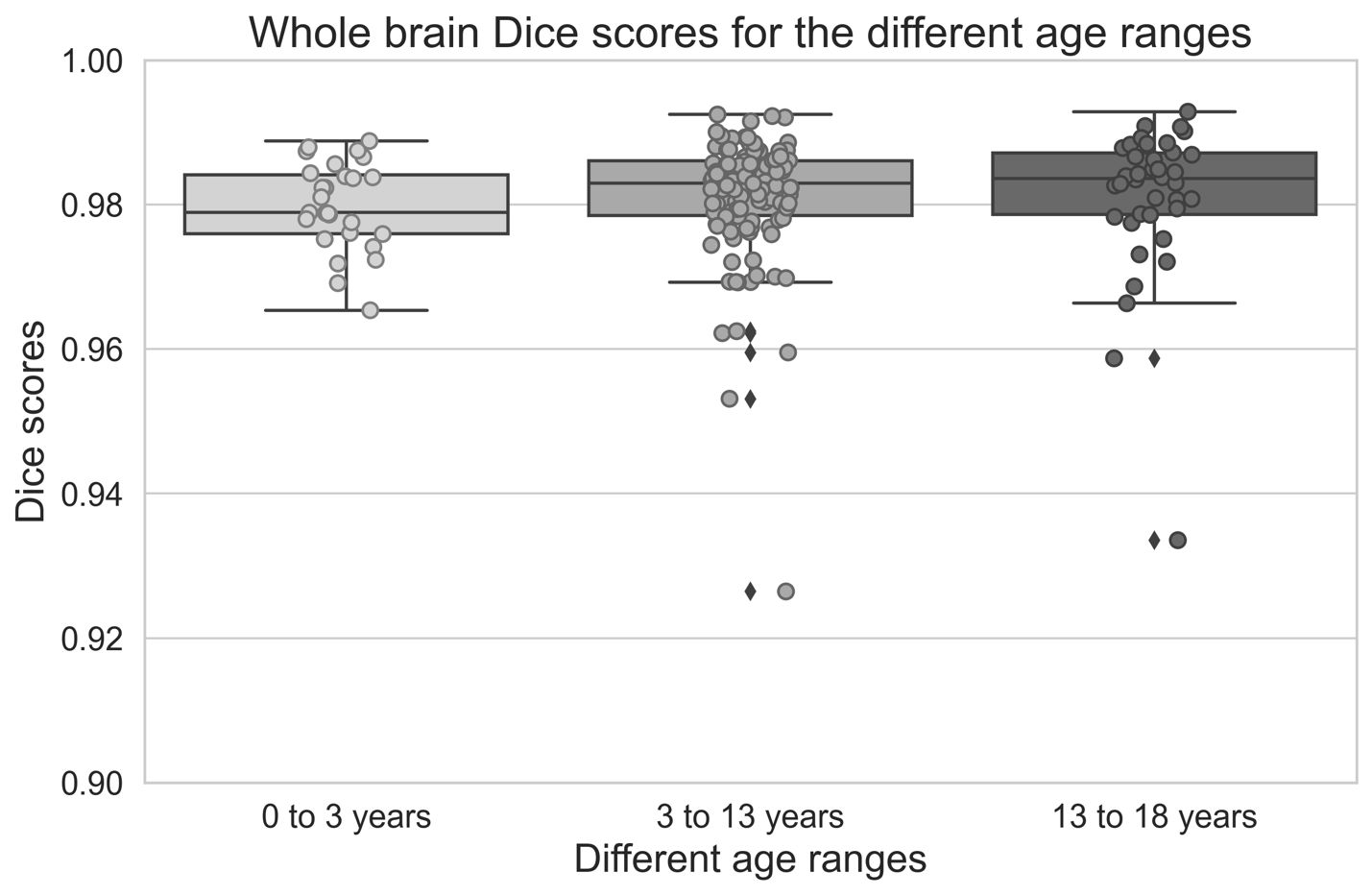

**Supplemental Figure 5.** Distribution of whole brain Dice scores using the multi-parametric skull-stripping model for different age ranges: 0 - 3 years, 3 - 13 years, and 13 - 18 years. There is no significant difference in the whole brain Dice scores between subjects from different age ranges (all p>0.05).

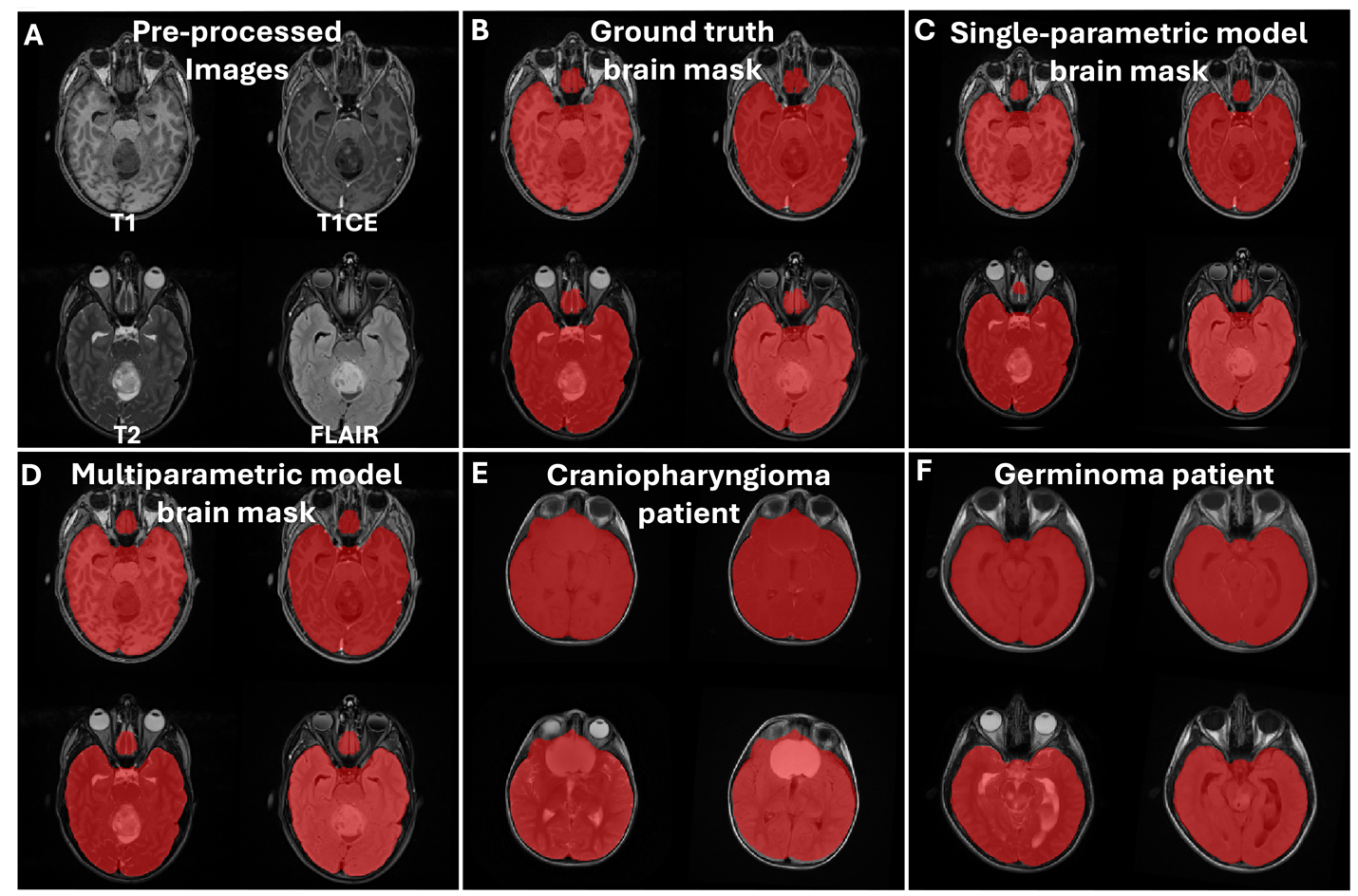

**Supplemental Figure 6.** Example pre-processed brain MR images (A) with overlaid ground truth brain mask segmentations (B), single-parametric skull-stripping model prediction (C), multi-parametric skull-stripping model prediction (D), along with example multi-parametric model predicted brain masks for a craniopharyngioma patient (E) and a germinoma patient (F). These images qualitatively demonstrate the superior performance of the two skull-stripping models.

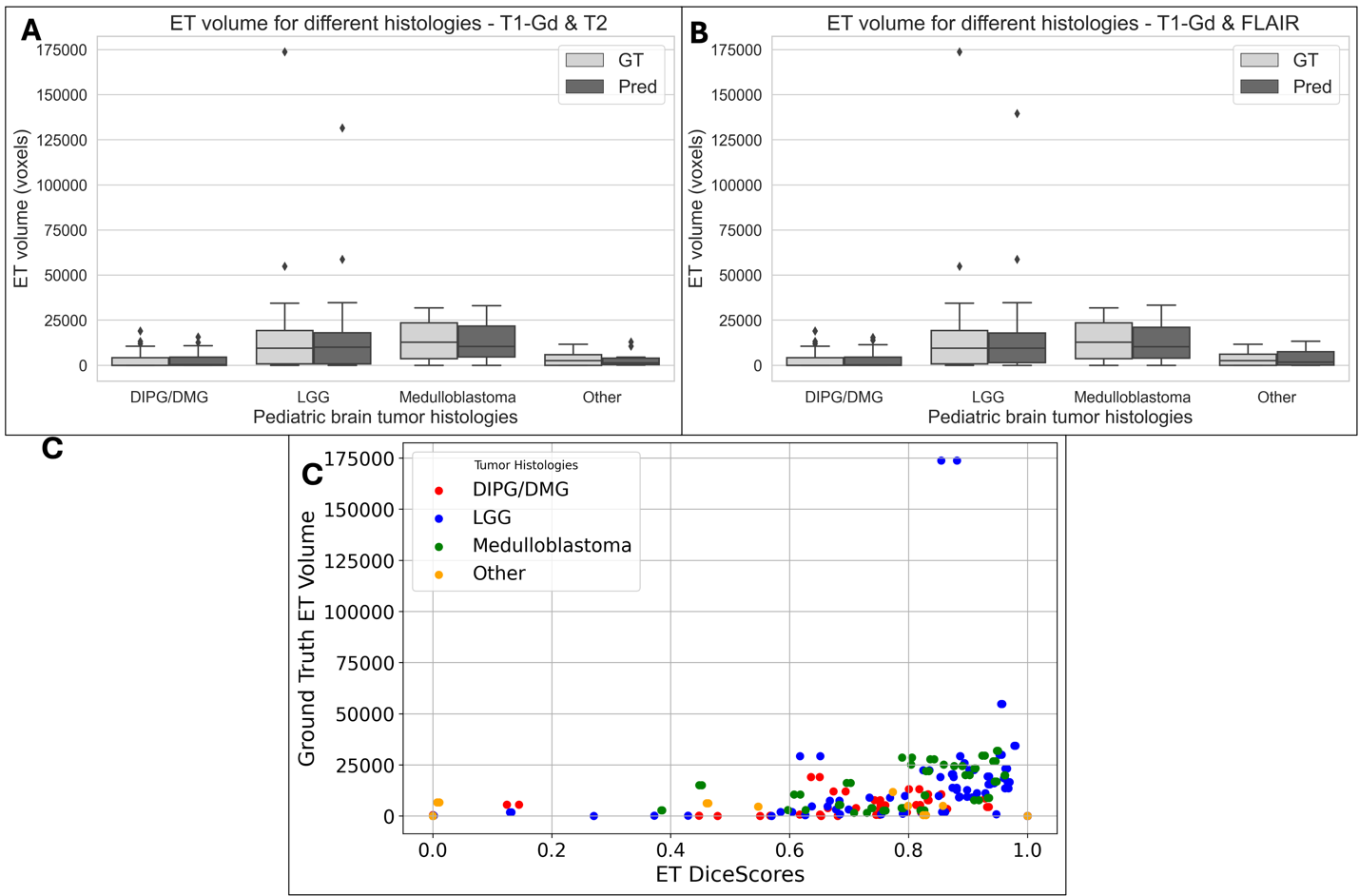

**Supplemental Figure 7.** Box plots showing the distribution of ET tumor volumes for ground truth and model-predicted segmentations with T1-Gd and T2 as inputs (A), and T1-Gd and FLAIR as inputs (B). Panel (C) shows the significant correlation between ET Dice scores and ground truth ET volumes.

**Supplemental Table 1.** Performance metrics for multi-parametric and single-parametric skull-stripping models (T1, T1-Gd, T2 and FLAIR inputs)– for both whole brain and sellar/suprasellar slices.

| Region / Metric | Dice scores  Mean ± sd (Median) | Sensitivity  Mean ± sd (Median) | 95% Hausdorff distance mm  Mean ± sd (Median) |
| --- | --- | --- | --- |
| **multi-parametric model – Whole brain** | | | |
| Brain Mask | 0.98±0.01 (0.98) | 0.98±0.02 (0.99) | 2.44±1.17 (2.24) |
| **multi-parametric model – Only sellar/suprasellar slices** | | | |
| Brain Mask | 0.98±0.01  (0.99) | 0.98±0.01  (0.99) | 1.06±0.32 (1.00) |
| **single-parametric model – Whole brain** | | | |
| T1 | 0.98±0.01 (0.98) | 0.98±0.02 (0.98) | 2.66±1.3 (2.24) |
| T1-Gd | 0.98±0.01 (0.981) | 0.98±0.01 (0.98) | 2.43±1.01 (2.24) |
| T2 | 0.98±0.01 (0.98) | 0.98±0.01 (0.98) | 2.56±1.04 (2.24) |
| FLAIR | 0.98±0.01 (0.98) | 0.98±0.02 (0.98) | 2.6±1.18 (2.24) |
| **single-parametric model – Only sellar/suprasellar slices** | | | |
| T1 | 0.98±0.01 (0.99) | 0.98±0.01 (0.98) | 1.12±0.40 (1.00) |
| T1-Gd | 0.98±0.01 (0.98) | 0.98±0.01 (0.99) | 1.09±0.27 (1.000) |
| T2 | 0.98±0.01 (0.98) | 0.98±0.02 (0.98) | 1.14±0.37 (1.00) |
| FLAIR | 0.98±0.02 (0.98) | 0.98±0.03 (0.98) | 1.15±0.59 (1.00) |

**Supplemental Table 2.** Performance metrics for whole tumor segmentation using skull-stripped versus non-skull-stripped images.

| **Patients with sellar/suprasellar tumors** | **WT Dice score (GT vs non-skull-stripped input)** | **WT Dice score (GT vs skull-stripped input)** |
| --- | --- | --- |
| **Dice scores**  **Mean ± sd (Median)** | **0.9±0.09(0.93)** | **0.89±0.1(0.93)** |
| **Sensitivity**  **Mean ± sd (Median)** | **0.86±0.11(0.89)** | **0.85±0.14(0.88)** |
| **95% Hausdorff distance mm**  **Mean ± sd (Median)** | **2.74±1.18(2.24)** | **3.01±1.35(2.53)** |

**Supplemental Table 3.** MRI scanner field strength and manufacturer information for the skull stripping and brain tumor segmentation model cohorts.

| **MRI scanner field strength and manufacturer information** | **Skull stripping model cohort** | **Brain tumor segmentation model cohort** |
| --- | --- | --- |
| **Total Patients** | **336** | **489** |
| **Scanner Magnetic Field Strength**  **0.7T**  **1.16T**  **1.5T**  **3T**  **Information unavailable** | **1**  **1**  **102**  **189**  **43** | **1**  **1**  **151**  **234**  **102** |
| **Scanner Manufacturer**  **Siemens**  **GE**  **Phillips**  **Toshiba** | **257**  **30**  **4**  **1** | **329**  **47**  **9**  **1** |
| **Hitachi**  **Information unavailable** | **1**  **43** | **1**  **102** |
